# Medical students’ perception of live lectures compared to video lectures in basic sciences medical education: a cross-sectional survey of medical colleges in Pakistan

**DOI:** 10.1101/2021.01.13.21249740

**Authors:** Maria Khan, Ali Bin Abdul Jabbar, Daniyal Ali Khan, Muhammad Abdullah Javed, Mohummad Hassan Raza Raja, Kanza Muzaffar

## Abstract

**Background:** Live lectures are commonly used in medical education, yet many students prefer video lectures instead. As different learning modalities may affect knowledge, it was necessary to explore medical students’ perspectives about the two learning modalities in Pakistan.

**Objectives:** This study aimed to explore and compare the medical students’ perspectives regarding live lectures and video lectures.

**Methods:** This cross-sectional study used an online questionnaire. This was distributed to medical students via internet platforms after institutional approval. Data were analyzed with SPSS version 23 using descriptive statistics.

**Results:** 585 students, from 11 medical colleges across six cities of Pakistan, were enrolled. 64.4% (n=377) of the students were females, while 34.0% (n=199) were males. The first years comprised 32.7% (n=191), second years, 29.2% (n=171), and third years, 38.1% (n=223) of the total. The commonest reason for attending live lectures was ‘they are compulsory’. The commonest reason for not attending was ‘poor teaching quality’. 5.0% (n=29) of 585 students reported live lectures and 51.8% (n=290 of 560) found video lectures to be ‘very helpful’ in concept clarification. 85.1% (n=258) of 303 students found video lectures more effective for learning. For 45.4% (n=254) of students, video lectures improved their grades a lot; more students used video lectures for exam preparation over the years. 50.6% (n=296) of students wanted video lectures to be compulsory, compared with 28.5% (n=167) for live lectures. The main improvement in live lectures was not using slides.

**Conclusion:** Medical students in Pakistan prefer video lectures over live lectures for learning and exam preparation. More students wanted video lectures to be compulsory in medical education. Several improvements have been suggested for live lectures.

**Summary box**
Section 1: What is already known on this topic?

- Medical students prefer video-based learning over attending live lectures.
- The choice of learning modality (video lectures versus live lectures) may affect knowledge and examination scores.
- Studies have not compared the effectiveness of commercial video lectures (created by any individual, not necessarily affiliated with a medical college, board examination preparation company, or independent person) to live lectures.

Section 2: What this study adds?

- Medical students prefer video lectures to live lectures in basic sciences medical education and most want video lectures to be compulsory in medical education.
- Medical students find video lectures more effective in concept clarification and use them more often in exam preparation and believe video lectures improve their grades.
- Medical students do not want (PowerPoint) slides to be used in live lectures.

## Introduction

Live lectures are a traditional means of imparting knowledge, where a lecturer delivers knowledge to students in a formal setting, with students present in the same room at the same time. There has been a decrease in the attendance of live lectures following the introduction of video-based learning.^1 2^ Likewise, medical students watch more video lectures in their second year compared to their first year.^3^ The causes of this are numerous: students use videos to facilitate learning in subjects they find challenging^4^; poor teaching quality; or early lecture timings.^1^ Video lectures are an increasingly common learning modality used by students. These are pre-recorded lectures that do not require physical interaction between the teacher and the student. They offer novel flexibilities, such as the self-determined speed of the lecture^5^ and accessing the lecture at home – which is where most students prefer to watch them.^6^

Despite their popularity, the effectiveness of video lectures as aids to learning remains debatable. According to some studies, students believe that videos help them perform better during their clinical rotations.^7 8^ However, other studies have found no significant difference between the effectiveness of video lectures compared to live lectures in imparting clinical knowledge.^9 10^ The two learning modalities may also directly impact academic performance. Video platforms have been shown to increase clinical knowledge, but they may not increase the knowledge of the actual subject.^11^ In addition, another study found that students who either attend lectures consistently or watch video lectures consistently score higher than those who mixed the two methods of study.^12^

A study conducted in Pakistan by Fatima et al shows that students prefer video-based learning over paper-based learning.^13^ However, most medical colleges in Pakistan conduct live lectures. Hence, it is likely that students in Pakistan, too, will be using video lectures to facilitate learning.

Despite this, students’ perceptions about video lectures have not been studied, with most studies focusing on ways to improve live lectures alone.^14 15^ In addition, studies have not explored the effectiveness of commercial video lectures, those that are created by any individual, not necessarily affiliated with a medical college, board examination preparing company or an independent person. Furthermore, current literature about the effectiveness of video lectures and live lectures are inconclusive and vary across countries, hence, it is important to conduct a local study to assess which modality is more commonly used by students and why, as it may affect their academic performance. Hence this study aims to assess medical students’ perceptions of video-based learning and live lectures and the effectiveness of each method in Pakistan. This will identify the shortcomings of the two learning modalities so they may be addressed.

## Materials and Methods

### Study Design and Setting

A cross-sectional survey was conducted in 11 medical colleges in six cities of Pakistan from October 2019 to December 2019.

### Sample size

A minimum sample size *(n)* of 363 has been calculated for the total population of N=6350, using a confidence interval of 95%, a margin of error of 5%, and a cooperation rate of 50%, using openepi.com, Version 3. Sample size *n* = [DEFF*Np(1-p)]/ [(d^2^/Z^2^_1-α/2_ *(N-1)+p*(1-p)] was used for this purpose. The sample size included all medical students enrolled in basic sciences. The subjects comprising basic sciences education are taught in the first three academic years in the medical colleges that were included in our survey; the only exception to this is the Aga Khan University where basic sciences are covered in the first two academic years.

### Sample selection

#### Inclusion criteria

All consenting medical students, of all genders, currently enrolled in their basic sciences years in the above-mentioned medical universities were included in this study.

#### Exclusion criteria

Individuals who did not provide consent (implied by not filling the questionnaire), or left the questionnaire incomplete, were excluded.

### Patient and Public Involvement

Not applicable.

### Operational definitions

Live lectures have been defined as lectures where the lecturer and the students are ‘face-to-face’, in the same room or lecture hall, in real-time.

Video lectures have been defined as pre-recorded lectures by any individual (not necessarily trained faculty of the relevant medical college), board examination preparation company, or independent persons.

### Exposures and outcomes

In this study, exposures included attending live lectures and watching video lectures, as defined above. Outcomes included live lecture attendance, the usefulness of learning modality in conceptual learning, preference of learning modality for examination preparation, changes in grades, and preference of learning modality as compulsory in medical education.

## Methods

The questionnaire for the study, “Should undergraduate lectures be compulsory?”^1^ was adapted with the permission of the authors. The questionnaire was distributed as an online Google Form and circulated via student data collectors.

## Ethics

Permission was sought and received from all participating medical colleges. This study received approval from the Ethics Review Committee at the Aga Khan University Medical College (ERC number 1771). All participants gave consent before participating in the study.

## Data Analysis

The data collected were tabulated and analyzed using IBM SPSS version 23 and Microsoft Excel 2010. Categorical data is represented as proportions, while simple descriptive variables are represented as percentages.

## Results

### Demographics

585 students from 11 medical colleges participated in this study. 33 students were enrolled in a medical college where lectures are not mandatory, while 552 students were enrolled in medical colleges where lectures are mandatory. 64.4% (n=377) of the total participants were females, while 34.0% (n=199) of the participants were males. 43.6% (n=255) of participants were day scholars, while 56.4% (n=330) of participants were residing in hostels. 32.7% (n=191) of participants were in their first year, 29.2% (n=171) in their second year, and 38.1% (n=223) in their third year of medical education. The results are shown in figure 1.

**Figure 1.**
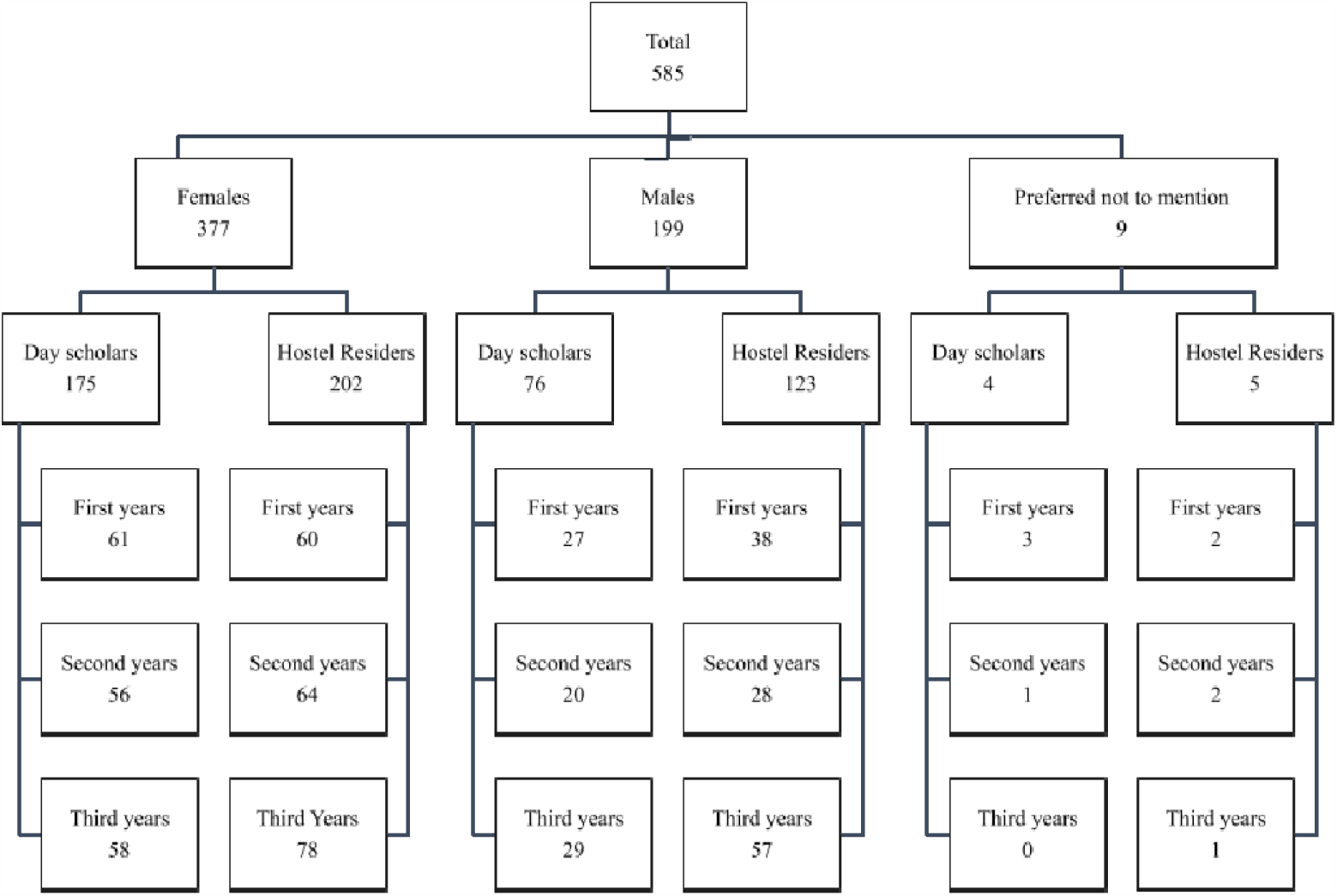
Demographics of participants from all medical colleges.

### Lecture Attendance

The minimum percentage of necessary attendance for medical colleges where lectures were mandatory was 75% for all medical colleges. From these medical colleges, out of 552 students, 28.4% (n=157) found it challenging to maintain attendance in their first year of medical education. In the second year of medical education, 49.5% (n=273) found it difficult to maintain attendance. Lastly, in the third year, 51.3% (n=283) found it difficult to maintain attendance above 75%. 22.1% (n=122) of students maintained an attendance of 90-100%, while 42.8% (n=236) of students had attendance in the range of 80-90% and 27.0% (n=149) in the range of 75-80%. Reasons for attendance and non-attendance of lectures are shown in table 1.

**Table 1.**
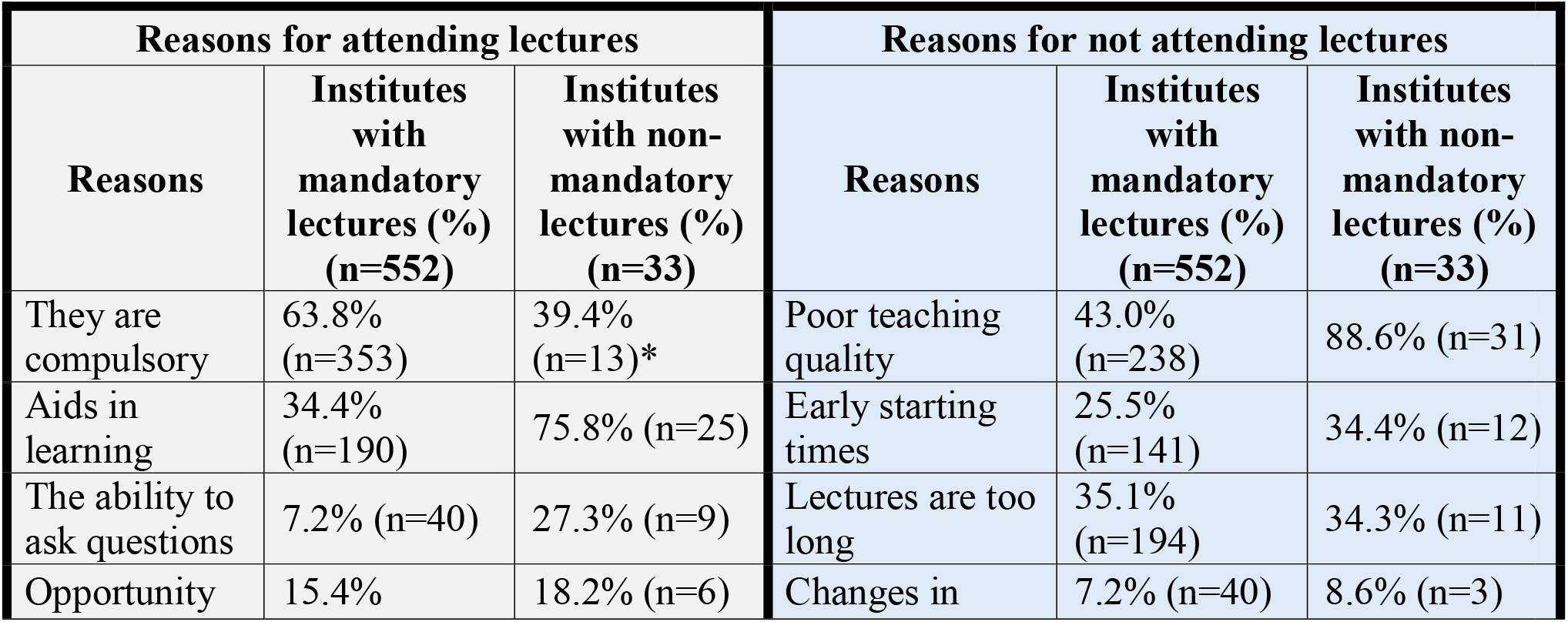
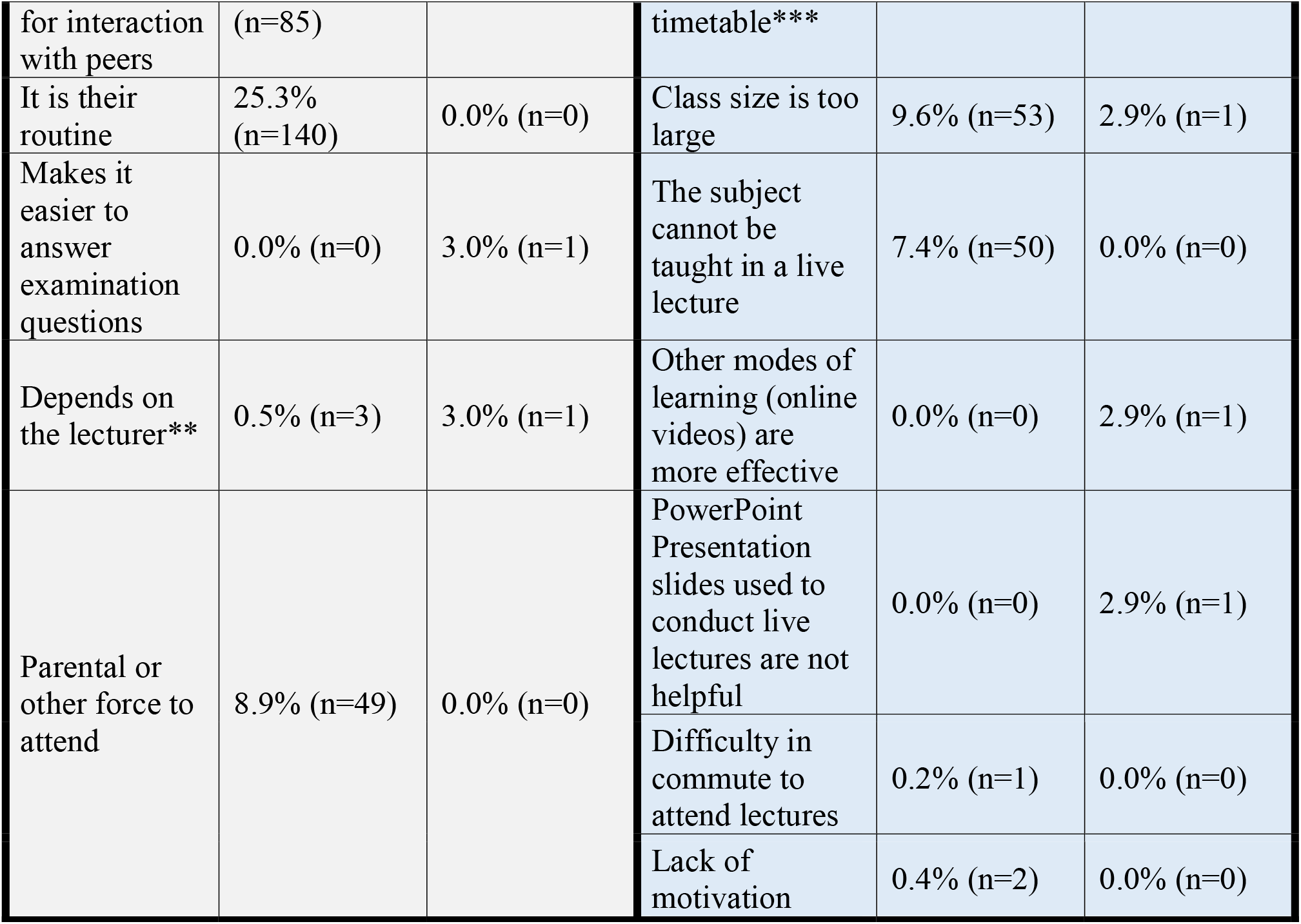
Reasons for attending and not attending lectures. *These students belong to the only university included in this study where lectures are not mandatory, however other sessions like problem-based learning, communication skills, longitudinal theme sessions, laboratory sessions and team-based learning are mandatory. **This implies how good the lecturer is. *** Changes in timetable implies preponing or postponing of lectures.

### Conceptual Learning

Students were asked how helpful live lectures or video lectures were in clarifying concepts (*with a p-value of <0.01, these results were statistically significant at a 99% confidence level). Out of 585 students, 5.0% (n=29) found live lectures ‘very helpful’, 28.5% (n=167) found them ‘somewhat helpful’, 36.1% (n=211) were neutral about it, 24.1% (n=141) chose ‘not very helpful’, and 6.3% (n=37) chose ‘not helpful at all’. On the other hand, out of 560 students who responded to this question, 51.8% (n=290) found video lectures ‘very helpful’, 42.5% (n=238) found them ‘somewhat helpful’, 5.4% (n=30) gave a neutral response, 0.4% (n=2) found them ‘not very helpful’, and none of them chose ‘not helpful at all’.

In the context of usefulness and effectiveness in learning, out of 303 students, 14.9% (n=45) of students preferred live lectures while 85.1% (n=258) preferred video lectures. Students were also asked if they found videos or video lectures useful for conceptual learning. 95.7% (n=536) of students responded ‘yes’, while 4.3% (n=24) of students responded ‘no’.

### Examination preparation

25.3% (n=148) of students reported that live lectures are helpful in exam preparation, 53.8% (n=315) reported that they are somewhat helpful, and 20.9% (n=122) reported that they are not helpful. The methods used by the participants in each year of their medical education are depicted in figures 2-4.

**Figure 2.**
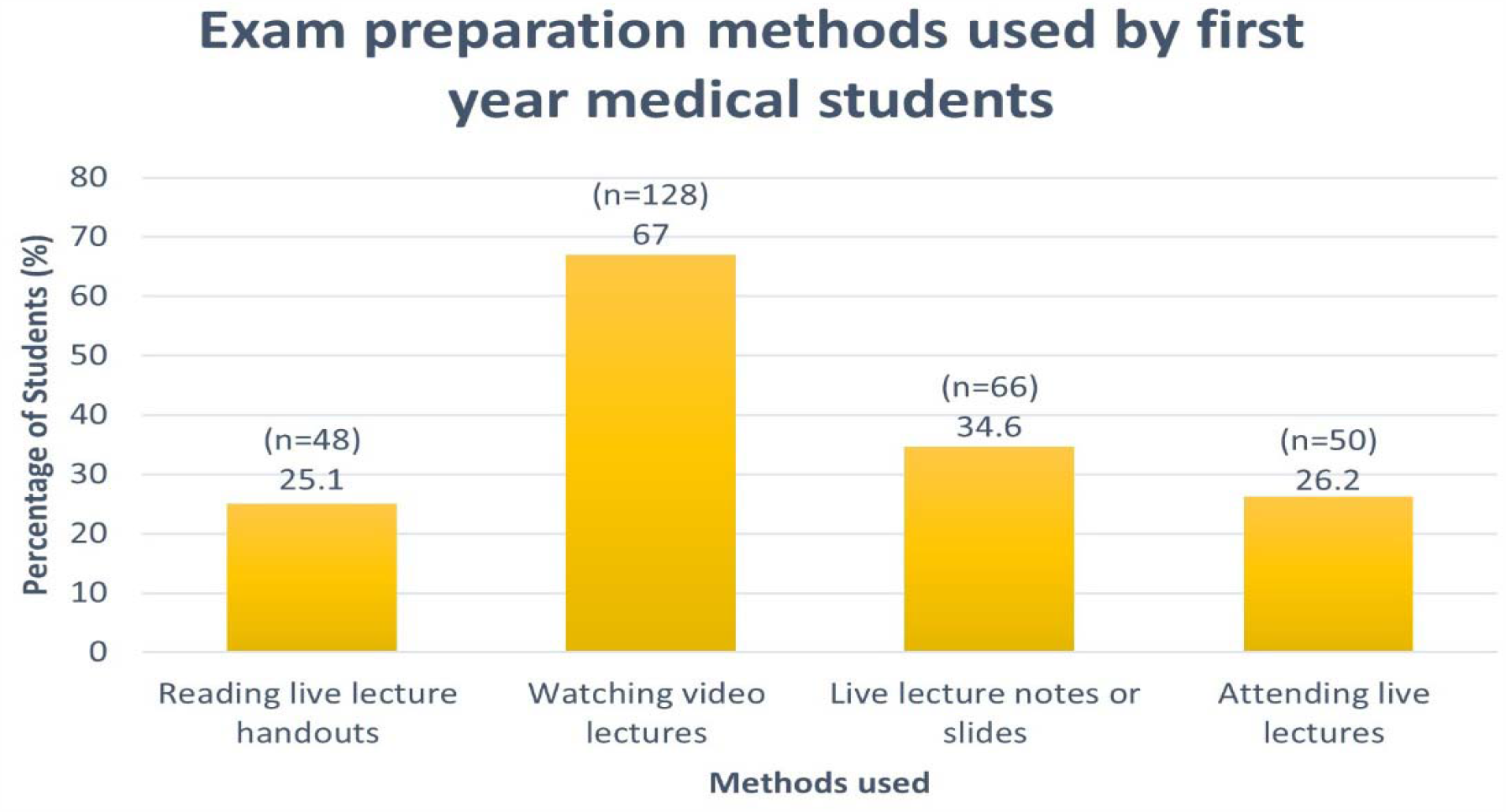
The material used by first-year medical students to prepare for their examinations. Participants could choose more than one learning modality in their response.

**Figure 3.**
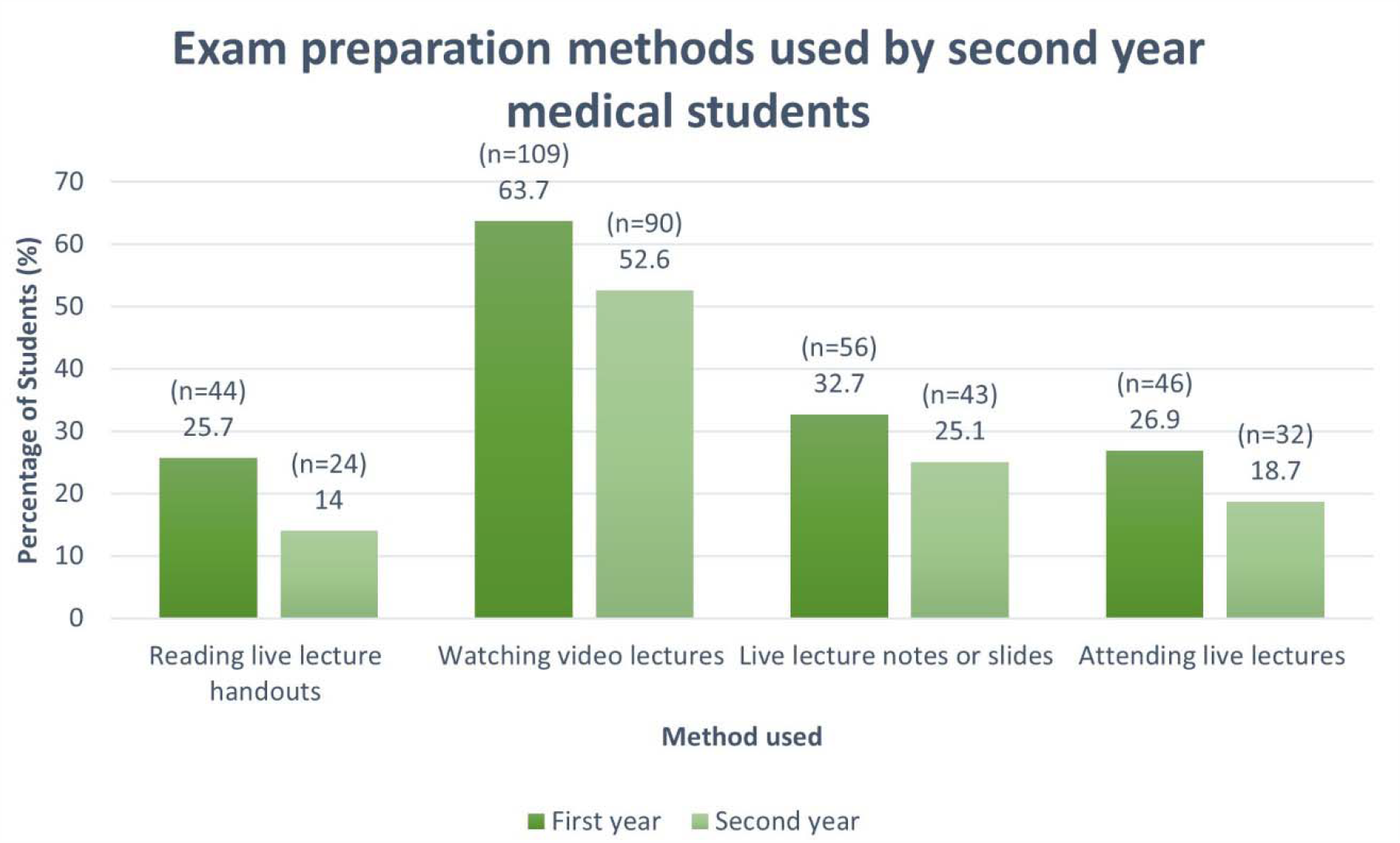
The material used by second-year medical students to prepare for their examinations in the first and second year of medical education. Answers could include more than one learning modality.

**Figure 4.**
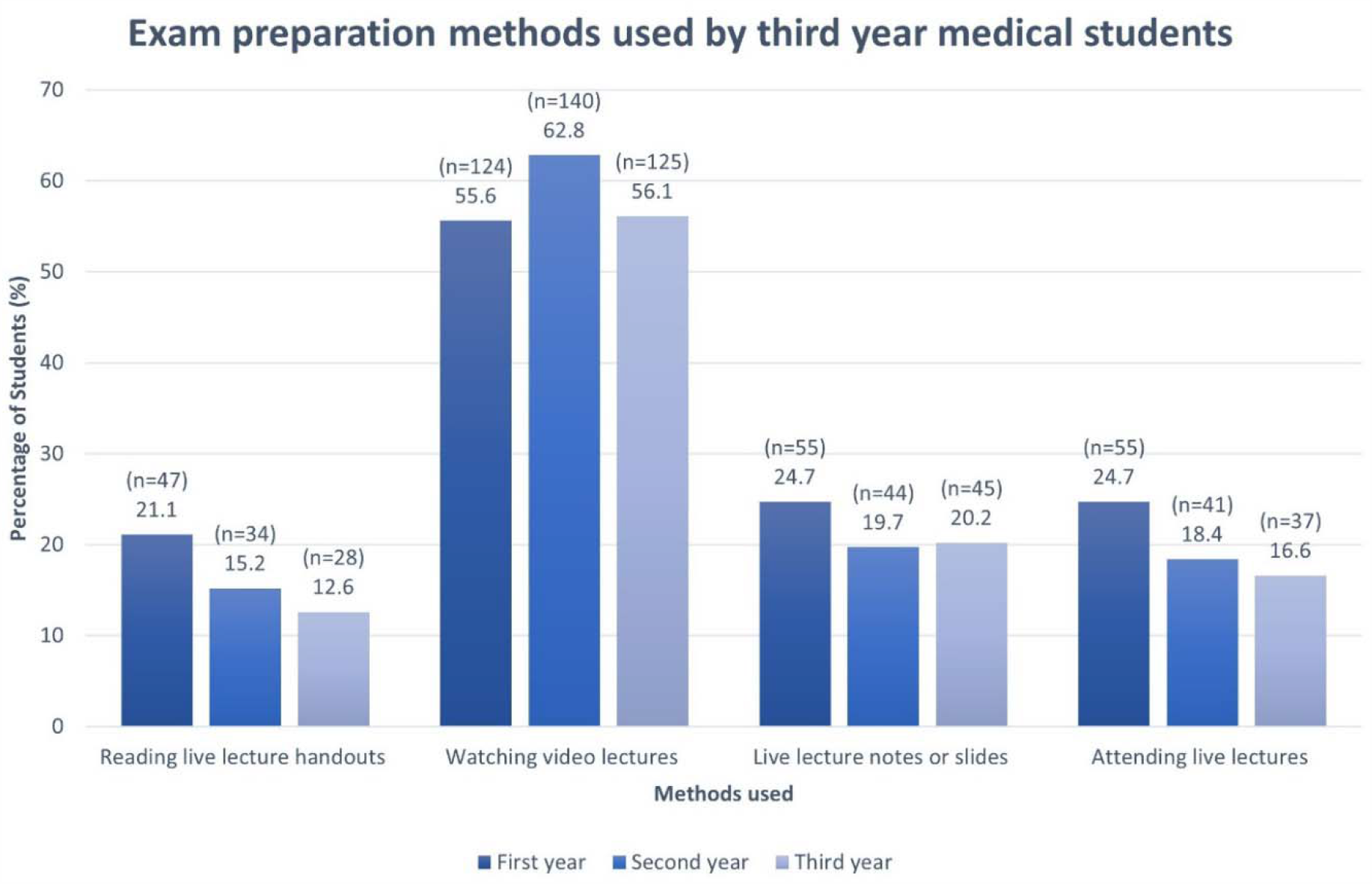
The material used by third-year medical students to prepare for their examinations in the first, second, and third year of medical education. Participants could choose more than one answer.

### Grades

Students were asked about the effect of video lectures on their grades and their examination performance. Grade changes were quantified as such: ‘improved a lot’ (>10% increase), ‘improved somewhat’ (5-10% increase), ‘stayed the same’ (+-5%), ‘gone down slightly’ (5-10% decrease), ‘gone down a lot’ (>10% decrease). Out of 560 students, 45.4% (n=254) said their grades ‘improved a lot’ and for 37.3% (n=209), they ‘improved somewhat’. Grades ‘stayed the same’ for 15.9% (n=89) of students while they ‘decreased slightly’ for 1.4% (n=8). None of the students selected ‘decreased a lot’. The examination performance was ‘much better’ for 50.0% (n=280), ‘slightly better’ for 41.8% (n=234), ‘same’ for 7.9% (n=44), ‘slightly worse’ for 0.4% (n=2), and ‘much worse’ for none of the students.

### Compulsory status of live lectures

Students were asked which learning modality should be compulsory in medical education. 28.5% (n=167) were in favor of live lectures, 50.6% (n=296) were in favor of video lectures, while 20.9% (n=122) chose the ‘neither/other’ modality option.

### Improvements

The students were asked to suggest improvements to live lectures and video lectures; each student could select multiple options. Improvements to live lectures suggested by students are summarized in table 2.

**Table 2.** Improvements to live lectures suggested by medical students.

| **Improvements to live lectures** | **Percentage of students (n)** |
| --- | --- |
| No slides used in lectures | 20.7% (n=34) |
| More interactive lectures | 17.7% (n=29) |
| Shorter lectures | 12.8% (n=21) |
| Teaching of more concepts | 12.2% (n=20) |
| Effective teaching | 11.0% (n=18) |
| More use of videos | 11.0% (n=18) |
| Non-mandatory lectures | 10.4% (n=17) |
| Adding case studies and clinical correlation | 6.7% (n=11) |
| Use of diagrams | 4.9% (n=8) |
| Teachers answering more questions | 4.3% (n=7) |
| Recap topics | 2.4% (n=4) |
| Teachers being more passionate about the subject | 1.8% (n=3) |
| Less usage of difficult terminology | 1.8% (n=3) |
| Content that helps answer examination questions | 1.8% (n=3) |
| Other | 8.5% (n=14) |
| Total | 100.0% (n=164) |

Only eight students suggested some improvements to video lectures. 50.0% (n=4) suggested that the video lectures should be available free of cost, 37.5% (n=3) suggested that they should have defined objectives, 25.0% (n=2) thought that the lectures should be concept-based, and 25.0% (n=2) suggested that they should be of shorter duration.

## Discussion

This study assessed the use of video lectures compared to live lectures as learning modalities by medical students. Our results were consistent with the literature, in that most students attended live lectures because they were compulsory,^1^ they helped in learning,^16^ or it was part of a student’s routine.^1 16^ Reasons such as asking questions and social interaction were less commonly reported in our study and the literature,^1 5 16^ however, they also play a part. More students from the cohort with non-mandatory lectures chose academic reasons for lecture attendance, such as live lectures being an aid to learning. This may be because, as the lectures were not mandatory, only students who were interested in learning attended them.^16^

The commonest reason for lecture absenteeism was ‘poor teaching quality’. One student mentioned, “I think it all comes down to who I actually want to listen to, there are teachers I’d prefer over Sketchy [video lectures], and vice versa. If the teacher delivers information in a clearer way, I wouldn’t have any problem sitting through a lecture.” Our results are in concordance with many other studies.^1 16-18^ Institutions should give importance to student feedback when evaluating teachers’ effectiveness and should train lecturers thoroughly. Live lectures usually last one-hour; shorter lecture times should be trialed. Class size also discouraged students from attending, with local colleges have more than 100 students per batch. Studies about the effect of class sizes on outcomes have been disputed,^19-21^ and more research is required in this area.

Students found video lectures more useful in clarifying concepts than live lectures. We assessed effectiveness using students’ self-reported concept clarification as well. This parameter is infrequently used in evaluating medical education. Most studies use test scores instead, yet results are inconclusive.^9 11 12^ Millis et al, when studying first-year medical students, found attendance/participation in the classroom to increased test scores.^22^ However, participation implied taking additional tests, which is exam practice. However, Fogleman et al found attendance affected scores in the National Board of Medical Examiner’s test.^23^ Yet, a randomized trial by Solomon et al and a study by Davis et al found no significant difference in test scores.^24 25^

More students used video lectures for exam preparation than live lectures or live lecture material in all three years of medical education, with the number increasing over the years. The use of live lectures or lecture material decreased over the years. Previous local studies found students to favor videos.^13^ In our study too, students voted for video lectures to be mandatory in medical education.

Students suggested that live lectures should not be delivered via slides (PowerPoint Presentations), because it was not helpful when lecturers “just read off slides”. Slides also deter a student from attending lectures.^16^ “In live lectures nowadays, the PowerPoint media is used, so some teachers just read out what is written [on] the slides without even trying to explain it. [If] they explain the slides, it would be much better and would increase the interests of students as well”, one student wrote.

The results of our study show that students wanted more interactive lectures: “I think teachers should try to use the board more instead of using slides all the time. It gets boring for the students and we mostly feel as though the teacher might only be reading from slides rather than actually teaching. This way we are less likely to actually listen to what the teacher is saying.”

This study questionnaire was circulated online, and the cohort of students who filled the questionnaire may not be representative of the entire student population. The questionnaire was self-administered, and some answers may have a bias. Additionally, ‘How do you prepare for exams?’ required recall by students in higher years. Despite this, this study was conducted in eleven medical colleges in three cities of Pakistan, giving a generalizable view on medical students’ perceptions. Most previous studies have compared live lectures and video lectures by the same lecturers. However, students are increasingly using video lectures as defined by our study, and this learning modality is not well studied.

Medical students in Pakistan prefer video lectures over live lectures. There are many aspects of live lectures that can be improved upon, including teaching quality, delivery style, and content. Such improvements may enhance students’ learning experience, increase live lecture attendance, and engage students more. Further studies may be done to assess students’ perceptions in more detail to be better able to target the various pitfalls of live lectures.

## Conclusion

The results of this study show that the majority of medical students in Pakistan prefer video lectures over live lectures. There are many aspects of live lectures that can be improved upon, including teaching quality, delivery style, and content. Such improvements may enhance students’ learning experience, increase live lecture attendance, and engage students more. Since this study was conducted in multiple institutes throughout Pakistan and the convenient sampling without any strict exclusion criteria, likely, the medical students in the study population who are not a part of this study had similar perceptions. We expect this study to have high external validity and recommend that another multi-institutional study should be conducted so that students’ perceptions can be assessed in more detail to be better able to target the various pitfalls of live lectures.

## Author contributions

Maria Khan planned and designed this study. Ali Bin Abdul Jabbar and Daniyal Ali Khan collected and analyzed the data. Muhammad Abdullah Javed and Muhammad Hassan Raza contributed to the questionnaire and editing the manuscript. Dr. Kanza Muzaffar supervised this study, with guidance at each stage. All authors contributed to and approved the final version of the manuscript to be published.

## Supporting information

STROBE checklist for cross-sectional studies

## Data Availability

We will share the data if requested to do so.
To request data, please send an email to

## Acknowledgments

We would like to acknowledge the valuable input from Dr. Kulsoom Ghias, Dr. Syeda Sadia Fatima, and Dr. Rehana Rehman regarding the adapted questionnaire.

## Data sharing

We will share the data if requested to do so.

## Copyright

I, the Submitting Author has the right to grant and does grant on behalf of all authors of the Work (as defined in the author license), an exclusive license and/or a non-exclusive license for contributions from authors who are: i) UK Crown employees; ii) where BMJ has agreed a CC-BY license shall apply, and/or iii) in accordance with the terms applicable for US Federal Government officers or employees acting as part of their official duties; on a worldwide, perpetual, irrevocable, royalty-free basis to BMJ Publishing Group Ltd (“BMJ”) its licensees.

## Transparency statement

The lead author (the manuscript’s guarantor) affirms that the manuscript is an honest, accurate, and transparent account of the study being reported; that no important aspects of the study have been omitted; and that any discrepancies from the study as planned (and, if relevant, registered) have been explained.

## Dissemination to participants and related patient and public communities

There are no plans to disseminate the results of the research to study participants since no identifiers were collected. Study results will be shared with the public via social media and conference presentations.

